# Quantitative detection of SARS-CoV-2 Omicron BA.1 and BA.2 variants in wastewater through allele-specific RT-qPCR

**DOI:** 10.1101/2021.12.21.21268077

**Authors:** Wei Lin Lee, Xiaoqiong Gu, Federica Armas, Fuqing Wu, Franciscus Chandra, Hongjie Chen, Amy Xiao, Mats Leifels, Feng Jun Desmond Chua, Germaine WC Kwok, Joey YR Tay, Claire YJ Lim, Janelle Thompson, Eric J Alm

## Abstract

On November 26, 2021, the World Health Organisation classified the B.1.1.529 SARS-CoV-2 variant as the Omicron variant of concern (VOC). Reports of higher transmissibility and potential immune evasion triggered flight bans and heightened health control measures across the world to stem its distribution. Wastewater-based surveillance has demonstrated to be a useful complement for community-based tracking of SARS-CoV-2 variants. Using design principles of our previous assays that detect VOCs (Alpha and Delta), here we report three allele-specific RT-qPCR assays that can quantitatively detect and discriminate the Omicron BA.1 and BA.2 variants in wastewater. The first assay targets the nine-nucleotide deletion at the L24-A27S of the spike protein for detection of BA.2. The second targets the six-nucleotide deletion at 69-70 of the spike protein for detection of the Omicron BA.1 variant, and the third targets the stretch of mutations from Q493R to Q498R for simultaneous detection of both Omicron BA.1 and BA.2. This method is open-sourced, can be implemented using commercially available RT-qPCR protocols, and would be an important tool for tracking the introduction and spread of the Omicron variants BA.1 and BA.2 in communities for informed public health responses.

## INTRODUCTION

The Coronavirus disease 2019 (COVID-19) was first detected late December 2019 and rapidly spread globally, leading to the WHO declaring it a global pandemic by March 2020 (WHO, 2020). The etiological cause of COVID-19 is the single-stranded RNA virus SARS-CoV-2 of the genus Betacoronavirus, a genus which also contains other human respiratory pathogens such as SARS-CoV and MERS-CoV (Pal et al., 2020). SARS-CoV-2 infection in the respiratory and gastrointestinal tract is mediated by the binding between viral spike protein (S) and human angiotensin-converting enzyme 2 (ACE-2) receptor (Zhou et al., 2020). Over the course of the COVID-19 pandemic, multiple SARS-CoV-2 variants of concern (VOCs) emerged due to genetic mutation of the viral genome. While many variants contain inconsequential mutations, some obtained mutations that confer higher fitness through increased transmissibility and the ability to elude medical countermeasures such as vaccines (Harvey et al., 2021). These variants are deemed by the WHO as variants of concerns due to their increased risk (CDC, 2021). To date, WHO has classified five variants as VOCs. These include Alpha (B.1.1.7), Beta (B.1.351), Gamma (P.1) and the Delta (B.1.617.2), a variant which emerged from India in October 2020 and quickly became the predominant global strain by mid-2021 (WHO, 2021a). Omicron (B.1.1.529), the fifth VOC and its descendent lineages were reported in multiple countries as early as November 2021. The Omicron variant draws attention due to its high number of mutations, with 26-32 of them in the spike protein (WHO, 2021b). The rapid expansion of the Omicron in the South African region and global distribution amidst early reports of higher transmissibility and immune escape compared to the Delta variant raised alarms for researchers and public health officials worldwide (Pulliam et al., 2021).

In light of the emergence of VOCs, surveillance efforts to track their introduction and spread in both naive and vaccinated populations become important to fight the pandemic and contain the spread of SARS-CoV-2. Currently, the most widely-used method for variant surveillance involves sequencing of clinical samples. However, the accessibility of genomic sequencing is limited and unsustainable due to the high capital and operational costs and the associated specialised infrastructure requirements (Gwinn et al., 2019). Furthermore, a significant fraction of confirmed cases needs to be sequenced to generate a meaningful dataset. A complementary method of surveillance that has been drawing a lot of attention during this pandemic is wastewater-based surveillance (WBS). WBS promises a robust, low-cost, real-time, and unbiased (i.e., indiscriminate of the manifestation of symptoms) snapshot of the entire populations within its sewershed/catchment range (Polo et al., 2020; Thompson et al., 2020). WBS has been shown to be effective, specific and comparatively sensitive at determining SARS-CoV-2 circulation trends during this COVID-19 pandemic across different countries (Medema et al., 2020; Randazzo et al., 2020; Wu et al., 2021). Further, WBS has been used to track the introduction and spread of VOC SARS-CoV-2 strains. Most of the efforts for variant tracking in wastewater rely on enriching and sequencing the environmental SARS-CoV-2 genome (Crits-Christoph et al., 2021; Fontenele et al., 2021; Napit et al., 2021). However, poor sensitivity towards low-frequency variants commonly found during the “introduction” of a VOC in a population and the lack of quantitative modelling of the data generated limits widespread application (Van Poelvoorde et al., 2021). RT-qPCR-based methods capable of targeting variant-specific mutants in the SARS-CoV-2 genome have been successfully demonstrated for variant identification in clinical samples (Wang et al., 2021) and we and others have recently adapted and validated use of such assays for quantification of multiple variants in wastewater samples (Graber et al., 2021; Lee et al., 2021a, 2021b, 2021c; Yaniv et al., 2021). RT-qPCR methods enable differentiation of specific variant-linked mutations and are more sensitive than sequence-based approaches, allowing for their quantitation, and providing readily interpretable results within hours and using widely available methods and workflows. However, designing assays for VOCs is a moving target since emerging VOCs may possess unique mutations that necessitate the development of new assays.

Our group has previously designed RT-qPCR-based methods for the detection of mutations associated with the Alpha variant and the Delta variant in wastewater (Lee et al., 2021a, 2021b, 2021c). The assays are designed based on principles of allele-specific (AS-) qPCR (Petruska et al., 1988; Wu et al., 1989). Here, we report three sets of AS RT-qPCR primers for the specific detection of the Omicron BA.1 and BA.2 variants - 1) assay targeting substitutions at loci Q493R to Q498R could be used to detect both Omicron BA.1 and BA.2 variants, 2) assay targeting deletions at loci H69-V70 could be used to detect for Omicron BA.1, 3) assay targeting deletions at loci 24-27 could be used to detect for Omicron BA.2. These assays can be run as a singleplex or as a multiplex (Lee et al., 2021a) for improved convenience.

## RESULTS AND DISCUSSION

In this work we report three sets of allele-specific (AS-) RT-qPCR primers that could be used to detect, quantify and differentiate the Omicron variants BA.1 and BA.2 in wastewater, based on mutations present in the spike gene in Omicron (**Table 1**). The assay targeting loci Q493R to Q498R could be used to detect both Omicron BA.1 and BA.2 variants, the assay targeting loci H69-V70 could be used to detect for Omicron BA.1 and the assay targeting loci 24-27 could be used to detect for Omicron BA.2. These genome sites are selected with reference to CoVariants (GISAID, 2021a). As compared to earlier versions of this work, we have updated the primer and probe sets to also include assays specific for the BA.2 subvariant. Initial validation with RNA templates demonstrate the sensitivity and specificity required for their utilisation in WBS assays.

**Table 1.** Three sets of AS RT-qPCR assays presented in this work. Assay BA2 24-27 targets the stretch of nine-nucleotide deletions at loci 24-27 which is unique to BA.2. Assay BA1 69-70 targets the stretch of six-nucleotide deletion at loci 69-70 present in BA.1. This six-nucleotide deletion is also present in the Alpha B.1.1.7 and Eta B.1.525 variants, however these variants are no longer common in circulation. Assay Om 493-498 detects nucleotide substitutions Q483R, Q498R, with and without G496S, and allows simultaneous detection of both Omicron BA.1 and BA.2.

| Target loci on the spike protein | 24-27 |  | 69-70 |  | 493-498 |  |
| --- | --- | --- | --- | --- | --- | --- |
| Assay | WT 24-27 | BA2 24-27 | WT 69-70 | BA1 69-70 | WT 493-498 | Om 493-498 |
| Detects | WT sequence (L24, P25, P26, A27) | 9 nucleotide deletion (L24-, P25-, P26-, A27S) | WT sequence (H69, V70) | 6 nucleotide deletion (H69-, V70-) | WT sequence (Q483, G496 Q498) | 2 - 3 nucleotide substitutions (Q483R, Q498R, with and without G496S) |
| 21K Omicron BA.1 | Yes |  |  | Yes |  | Yes |
| 21L Omicron BA.2 |  | Yes | Yes |  |  | Yes |
| 21A Delta B.1.617.2 | Yes |  | Yes |  | Yes |  |

### Specificity and cross-reactivity of AS RT-qPCR primers against Omicron BA.1 or BA.2 and WT RNA

We validated the respective assays against synthetic full length WT and Omicron BA.1 or BA.2 RNA genome constructs (**Figure 1**). All mutant-specific assays do not cross-react with WT RNA at and below 10^3^ copies, conferring sufficient specificity for determining wastewater titers of the Omicron variants, given that the number of copies of SARS-CoV-2 RNA in each reaction containing RNA template from wastewater has been reported typically below 10^3^ (Duvallet et al., 2021; Wu et al., 2022, 2021, 2020). The amplification efficiencies of all the assays for their target RNA are above 85%.

**Figure 1.**
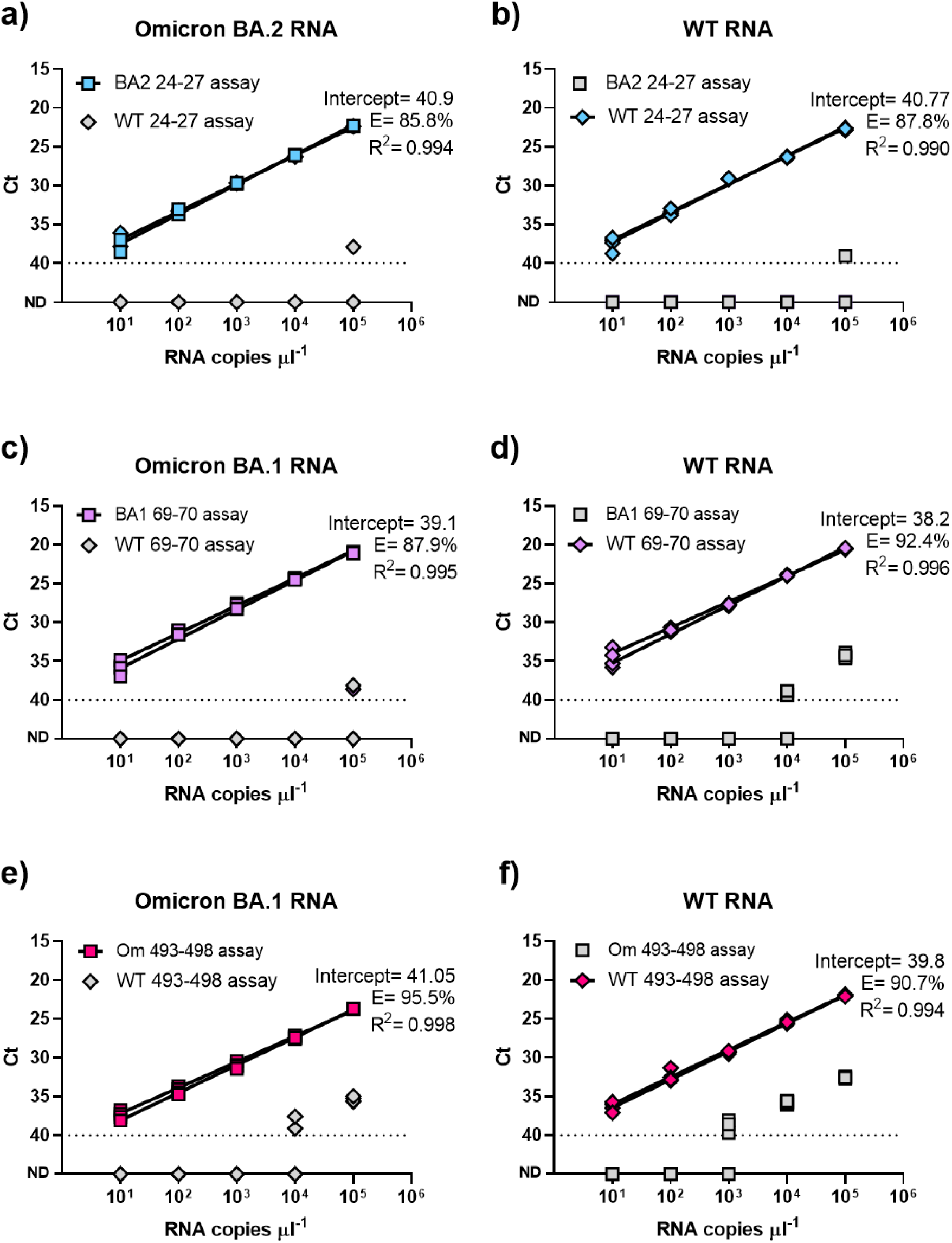
Specificity and cross-reactivity of AS RT-qPCR primers against full length WT and Omicron RNA. Squares represent tests of mutant-specific primers to Omicron BA.1 or BA.2 RNA, and diamonds represent tests of WT-specific primers to WT RNA (represented by Delta RNA). Grey diamonds and squares denote tests against RNA of the opposite genotype. The presented data reflect two sets of independent measurements taken on different days.

Using cycle thresholds (Ct) as a proxy for the sensitivity of the assays (**Figure 2**), we compared Ct values of the AS RT-qPCR assays developed, to those of the U.S. CDC N1 assay across a 10,000-fold difference in target RNA concentrations. WT and BA.1 assays targeting 69-70 and BA.2 assays targeting 24-27 had comparable sensitivities to the N1 assay while the rest of the assays had slightly lower sensitivities compared to the N1 assay. Nonetheless, all assays were able to reproducibly and robustly detect as little as 10 copies of target RNA.

**Figure 2.**
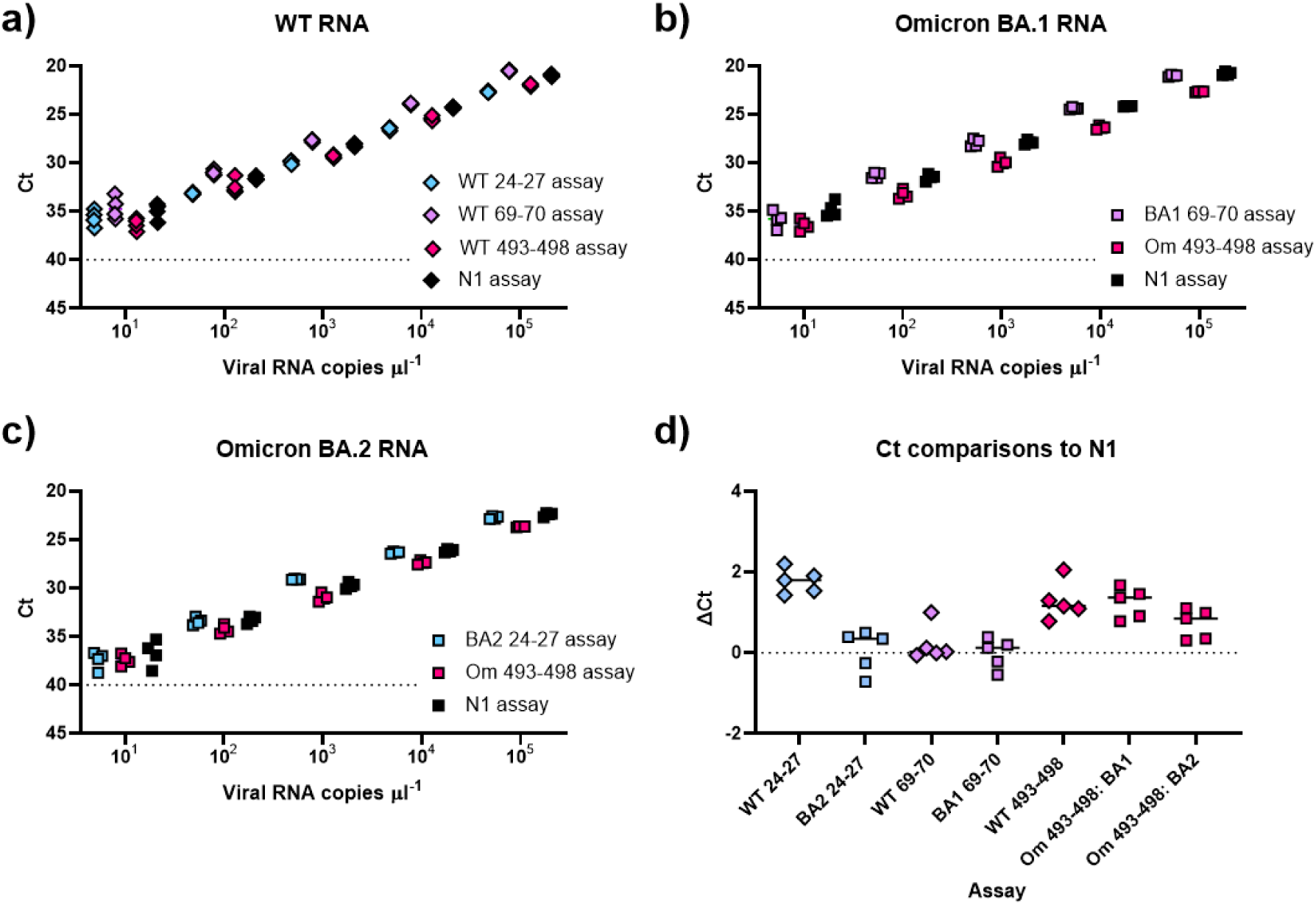
Cycle threshold (Ct) values for the AS RT-qPCR primers targeting 24-27, 69-70 and 493-498 in comparison to the US CDC N1 assay. AS RT-qPCR assays (blue, purple and pink) in comparison to the US CDC N1 assay (black). **a**) Diamonds denote tests against WT RNA (represented by Delta RNA), **b**) squares represent tests against Omicron BA.1 or **c**) BA.2 RNA. Assays were tested with ten-fold dilutions of their respective full-length synthetic Omicron SARS-CoV-2 BA.1 or BA.2 RNA or WT SARS-CoV-2 RNA in nuclease-free water. The data shown reflect two sets of independent measurements taken on different days. **d**) The difference in Ct of the respective assays in comparison to the N1 assay.

## DISCUSSION

The study presented here proposes three AS RT-qPCR assays that have been developed for specific detection and quantitation of the SARS-CoV-2 Omicron variant BA.1 and BA.2 in wastewater samples. The assay targeting loci 69-70 could be used to detect for Omicron BA.1, the assay targeting loci 24-27 for detecting the Omicron BA.2 and the assay targeting loci 493-498 for simultaneous detection of both Omicron BA.1 and BA.2 variants in complex wastewater matrices. All the assays reported in this work are highly specific and did not cross-react with RNA of their respective opposite genotypes in concentrations of up to 10^3^. All assays showed good amplification efficiencies and sufficient sensitivity and specificity to allow for the detection of the Omicron variants in wastewater.

The assay covering locus 69-70 was previously developed for the detection of the SARS-CoV-2 Alpha variant (Lee et al., 2021b,c). Due to co-evolution, the deletion at locus 69-70 can be found in the Alpha and the Eta variants as well as the more recent Omicron BA.1 VOC. While there may be a theoretical ambiguity if a positive read out with this assay would signify the Alpha, Eta or the Omicron BA.1 variant, but as of late Dec 2021, less than 0.2% of newly reported sequences is attributed to the Alpha and the Eta variants (GISAID, 2021b).

While not yet demonstrated in this work, the three assays reported in this work can be combined into a multiplex for greater convenience of handling. Variant detection requires performing only the variant-specific assays, of which the assays targeting the Omicron variant (BA.2 24-27 assay, BA.1 69-70 assay and Omicron 493-498 assay) can be combined into a reaction as a multiplex. Likewise the three WT-specific assays (WT 24-27 assay, WT 69-70 assay and WT 493-498 assay) can be combined as another multiplex. Potential multiplex combinations are shown in **Table 4**.

While variant detection only requires performing the reaction that targets the mutated locus in the SARS-CoV-2 variant of concern, we present assays that target both the WT and the mutant at the same loci. This way, quantification of both WT and mutant loci in wastewater allows one to determine the proportion of WT to the variant sequence at the target loci, if desired. In the first version of this study, Omicron variant RNA was not yet commercially available in our laboratory. Therefore, the WT assay was utilised to approximate the performance of the Omicron assay. In the subsequent second version, we updated the workflow to include a validation of the 69-70 and 493-498 assays using a full length synthetic Omicron BA.1 RNA construct. Following the emergence of the Omicron BA.2 variant, we further updated this study to include assays targeting loci 24-27 for specific detection of the Omicron BA.2 variant. We are sharing these assays and their initial validations for the benefit of the wider wastewater surveillance community.

## MATERIALS AND METHODS

### Assay design

We designed AS RT-qPCR reactions to detect a stretch of mutations in the SARS-CoV-2 spike gene. Primers and probes were designed following our previous work (Lee et al., 2021) and using the Integrated DNA Technologies (IDT)’s PrimerQuest Tool. Target mutations were placed near the 3’ end of the forward primer. All primers were designed to have a melting temperature in the range of 59–65°C and the probes in the range of 64–72°C. Probes were designed to anneal to the same strand as the allele-specific primer, with the probe as close to the 3′-end of the Allele Specific (AS) primers as possible. Guanines are avoided at the 5′-end of the probe. WT primers are designed to bind to all non-Omicron sequences. The probe for 493-498 is designed with mixed bases at two positions to enable binding to both WT (non-Omicron) and Omicron sequences. The primer F-WT-24-27 is designed with mixed bases at one position to enable binding to more WT (non-BA.2) sequences. All primers and probes were purchased from IDT (**Table 2**).

**Table 2.**
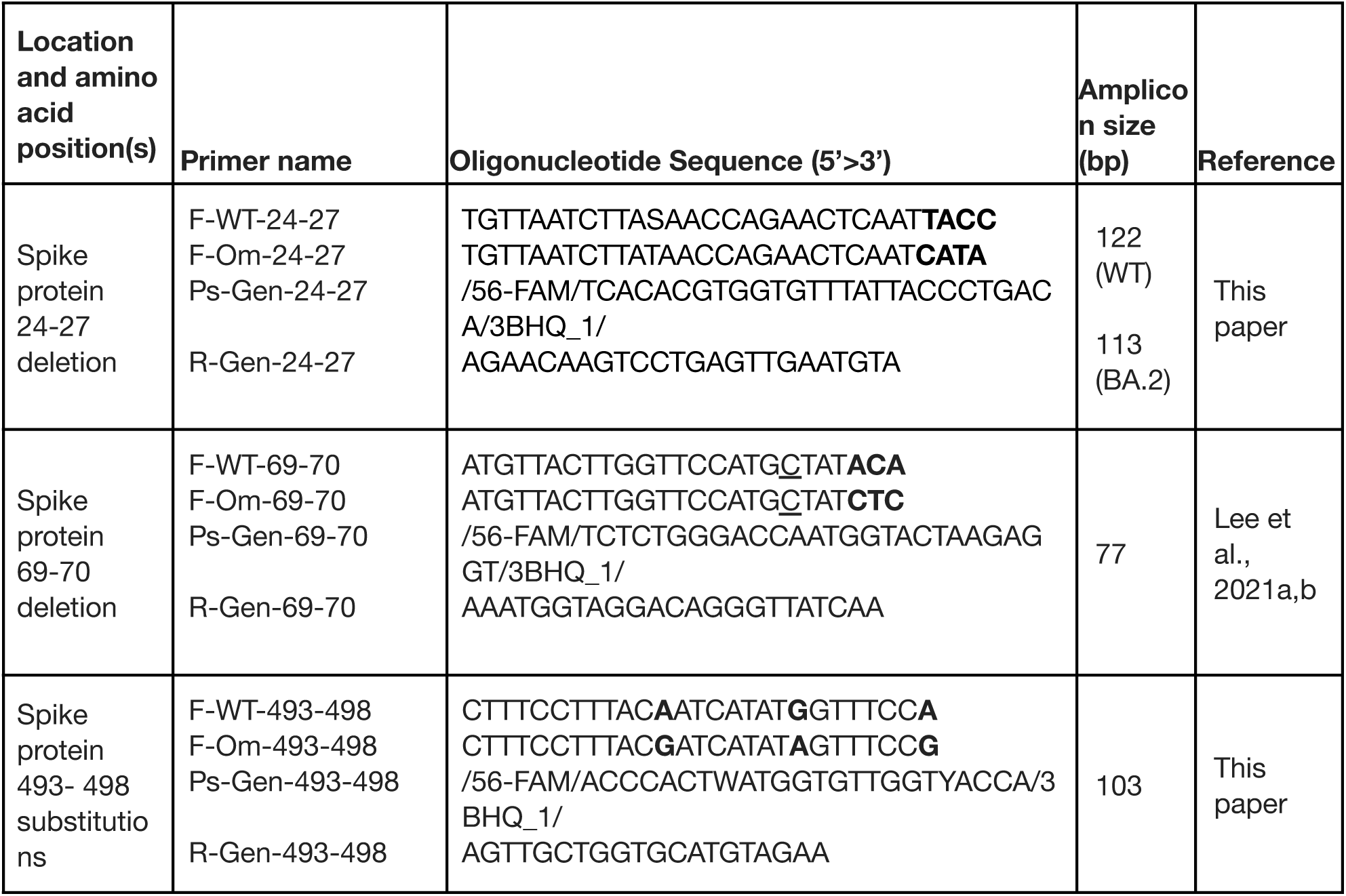
AS RT-qPCR primer sequences. Allele-specific nucleotides are marked bold. Gen denotes general (common primers), and are used in both WT and omicron-specific reactions. Om denotes Omicron, and is used in Omicron-specific reactions. WT denotes wild type, and is used for WT (non-Omicron)-specific reactions. If the assays were to be multiplexes, the probes should be synthesised with a different fluorophore i.e. HEX or Cy5. The assay for the locus 69-70 has been developed in an earlier study on Alpha variant quantification (Lee et al., 2021a, 2021b). The presence of the A67V mutation present as a C to T substitution in Omicron which is absent in Alpha, did not affect the performance of the 69-70 assay against Omicron RNA. Nucleotide underlined may be changed to a T to account for the A67V that is present in Omicron but not in Alpha. Retaining it as a C does not affect assay performance. While F-Om-493-498 is designed based on Q483R, G496S and Q498R, it does not discriminate between BA.1 (Q483R, G496S and Q498R) and BA.2 (Q483R and Q498R).

### RNA Standards and their quantification by RT-ddPCR

Twist synthetic SARS-CoV-2 RNA control 23 (Delta, B.1.617.2) was used as the WT RNA standard, as it contains the wild-type (WT) sequence at the three targeted mutant loci. Twist synthetic SARS-CoV-2 RNA control 48 (B.1.1.529/BA.1) and 50 (B.1.1.529/BA.2) were used as the RNA standard for Omicron BA.1 and BA.2 respectively. RNA standards were prepared as single-use aliquots. Controls 23, 48 and 50 were quantified by digital droplet RT-PCR (dd RT-PCR) to be 3.96 × 10^5^ copies/μL, 4.55 × 10^5^ copies/μL and 4.21 × 10^5^ copies/μL respectively. Quantification was performed using One-Step RT-ddPCR Advanced Kit for Probes #1864022 (Bio-Rad) following manufacturer’s recommendations.

### Analysis of assays against RNA standards by RT-qPCR

AS RT-qPCR was performed using the Taqman Virus 1-Step master mix (Thermofisher #4444434) with technical duplicates, at a final volume of 10 µL, according to the manufacturer’s recommendations. A single reverse primer and probe was used with each allele-specific forward primer (**Table 3**). The final concentration of the AS RT-qPCR primers were 500 nM, probe at 200 nM, with 1 µL of template. No template controls were included for each assay and none of them amplified. The reactions are setup using electronic pipettes (Eppendorf) and performed on a Bio-Rad CFX384 real-time PCR instrument under the following conditions, 5 min at 50 °C and 20 s at 95 °C, followed by 45 cycles of 3 s at 95 °C and 30 s at 60 °C.

**Table 3.**
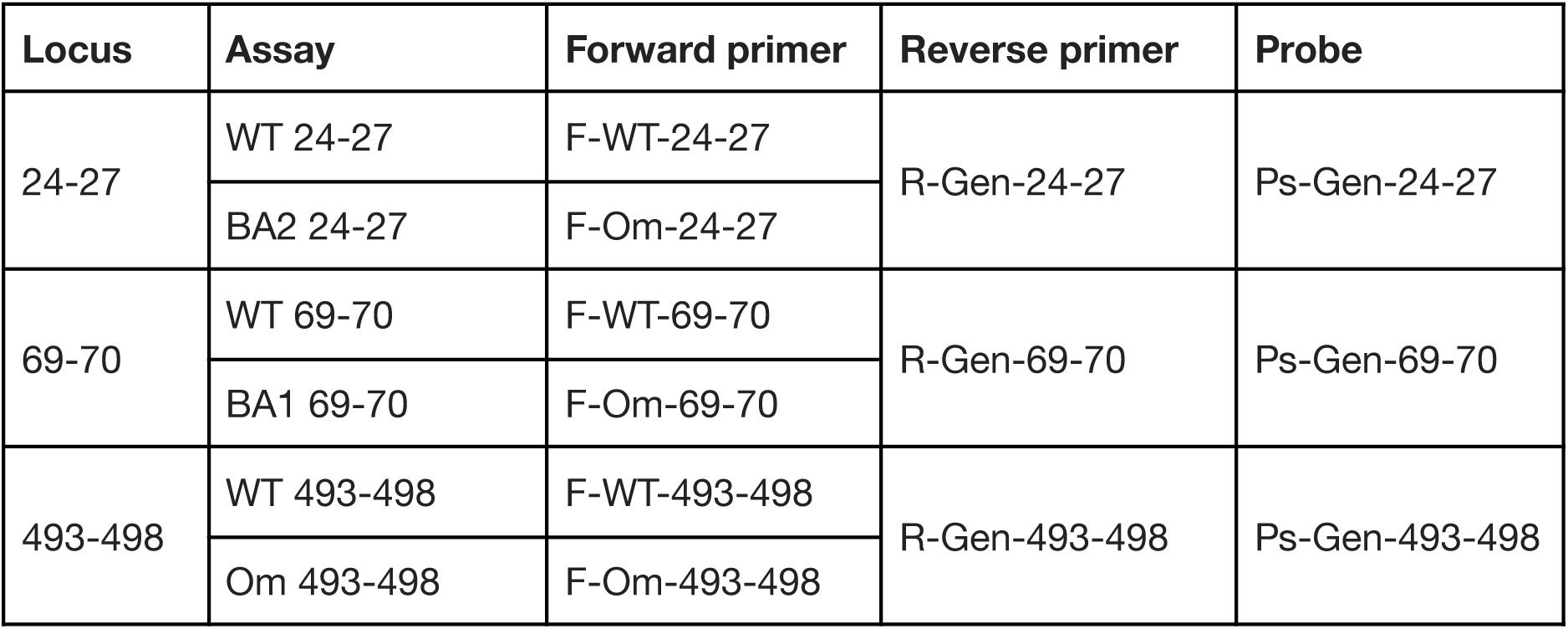
Primers and probes in each assay.

| Locus | Assay | Forward primer | Reverse primer | Probe |
| --- | --- | --- | --- | --- |
| 24-27 | WT 24-27 | F-WT-24-27 | R-Gen-24-27 | Ps-Gen-24-27 |
|  | BA2 24-27 | F-Om-24-27 |  |  |
| 69-70 | WT 69-70 | F-WT-69-70 | R-Gen-69-70 | Ps-Gen-69-70 |
|  | BA1 69-70 | F-Om-69-70 |  |  |
| 493-498 | WT 493-498 | F-WT-493-498 | R-Gen-493-498 | Ps-Gen-493-498 |
|  | Om 493-498 | F-Om-493-498 |  |  |

**Table 4.** Multiplex assay set up.

| **WT multiplex** | **Mutant multiplex** |
| --- | --- |
| F-WT-493-498<br>R-Gen-24-27<br>Ps-Gen-24-27 (Fam) | F-Om-24-27<br>R-Gen-24-27<br>Ps-Gen-24-27 (Fam) |
| F-WT-69-70<br>R-Gen-69-70<br>Ps-Gen-69-70 (Hex) | F-Om-69-70<br>R-Gen-69-70<br>Ps-Gen-69-70 (Hex) |
| F-WT-493-498<br>R-Gen-493-498<br>Ps-Gen-493-498 (Cy5) | F-Om-493-498<br>R-Gen-493-498<br>Ps-Gen-493-498 (Cy5) |

### Data analysis

qPCR data was analysed using Microsoft Excel and Graphpad Prism. Graphs were presented using Graphpad Prism.

## Data Availability

Source data will be made available upon request.

## Contributions

EJA and JT conceptualized the project. WLL designed the experiments. WLL, XG, FA, FC, HC, HC, FW, AX, ML, FJDC and GWCK analyzed the data. WLL, JYRT and CYJL performed experiments. All authors contributed to writing the manuscript. WLL, JT and EJA supervised the project. All authors read and approved the manuscript.

## Acknowledgements

We thank members of the Biobot Analytics, Inc team for helpful discussions.

## REFERENCES

CDC, 2021. SARS-CoV-2 Variant Classifications and Definitions [WWW Document]. URL https://www.cdc.gov/coronavirus/2019-ncov/variants/variant-info.html (accessed 7.9.21).

Crits-Christoph, A., Kantor, R.S., Olm, M.R., Whitney, O.N., Al-Shayeb, B., Lou, Y.C., Flamholz, A., Kennedy, L.C., Greenwald, H., Hinkle, A., Hetzel, J., Spitzer, S., Koble, J., Tan, A., Hyde, F., Schroth, G., Kuersten, S., Banfield, J.F., Nelson, K.L., 2021. Genome Sequencing of Sewage Detects Regionally Prevalent SARS-CoV-2 Variants. MBio 12, e02703–20. https://doi.org/10.1128/mBio.02703-20

Duvallet, C., Wu, F., McElroy, K.A., Imakaev, M., Endo, N., Xiao, A., Zhang, J., Floyd-O’Sullivan, R., Powell, M.M., Mendola, S., Wilson, S.T., Cruz, F., Melman, T., Sathyanarayana, C.L., Olesen, S.W., Erickson, T.B., Ghaeli, N., Chai, P., Alm, E., Matus, M., 2021. Nationwide trends in COVID-19 cases and SARS-CoV-2 wastewater concentrations in the United States. medRxiv 2021.09.08.21263283. https://doi.org/10.1101/2021.09.08.21263283

Fontenele, R.S., Kraberger, S., Hadfield, J., Driver, E.M., Bowes, D., Holland, L.A., Faleye, T.O.C., Adhikari, S., Kumar, R., Inchausti, R., Holmes, W.K., Deitrick, S., Brown, P., Duty, D., Smith, T., Bhatnagar, A., Yeager, R.A., Holm, R.H., von Reitzenstein, N.H., Wheeler, E., Dixon, K., Constantine, T., Wilson, M.A., Lim, E.S., Jiang, X., Halden, R.U., Scotch, M., Varsani, A., 2021. High-throughput sequencing of SARS-CoV-2 in wastewater provides insights into circulating variants. medRxiv 2021.01.22.21250320. https://doi.org/10.1101/2021.01.22.21250320

Graber, T.E., Mercier, É., Bhatnagar, K., Fuzzen, M., D’Aoust, P.M., Hoang, H.-D., Tian, X., Towhid, S.T., Plaza-Diaz, J., Eid, W., Alain, T., Butler, A., Goodridge, L., Servos, M., Delatolla, R., 2021. Near real-time determination of B.1.1.7 in proportion to total SARS-CoV-2 viral load in wastewater using an allele-specific primer extension PCR strategy. Water Res. 205, 117681. https://doi.org/10.1016/j.watres.2021.117681

GISAID, 2021a. Shared mutations [WWW Document]. URL https://covariants.org/shared-mutations (accessed 12.19.21).

GISAID, 2021b. Tracking of variants [WWW Document]. URL https://www.gisaid.org/hcov19-variants/ (accessed 21.19.21).

Gwinn, M., MacCannell, D., Armstrong, G.L., 2019. Next-Generation Sequencing of Infectious Pathogens. JAMA 321, 893–894. https://doi.org/10.1001/jama.2018.21669

Harvey, W.T., Carabelli, A.M., Jackson, B., Gupta, R.K., Thomson, E.C., Harrison, E.M., Ludden, C., Reeve, R., Rambaut, A., Peacock, S.J., Robertson, D.L., Consortium, C.-19 G.U.K. (COG-U., 2021. SARS-CoV-2 variants, spike mutations and immune escape. Nat. Rev. Microbiol. 19, 409–424. https://doi.org/10.1038/s41579-021-00573-0

Lee, W.L., Gu, X., Armas, F., Chandra, F., Chen, H., Wu, F., Leifels, M., Xiao, A., Desmond Chua, F.J., Kwok, G.W.C., Jolly, S., Lim, C.Y.J., Thompson, J., Alm, E.J., 2021a. Quantitative SARS-CoV-2 tracking of variants Delta, Delta plus, Kappa and Beta in wastewater by allele-specific RT-qPCR. medRxiv 2021.08.03.21261298. https://doi.org/10.1101/2021.08.03.21261298

Lee, W.L., Imakaev, M., Armas, F., McElroy, K.A., Gu, X., Duvallet, C., Chandra, F., Chen, H., Leifels, M., Mendola, S., Floyd-O’Sullivan, R., Powell, M.M., Wilson, S.T., Berge, K.L.J., Lim, C.Y.J., Wu, F., Xiao, A., Moniz, K., Ghaeli, N., Matus, M., Thompson, J., Alm, E.J., 2021b. Quantitative SARS-CoV-2 Alpha Variant B.1.1.7 Tracking in Wastewater by Allele-Specific RT-qPCR. Environ. Sci. Technol. Lett. https://doi.org/10.1021/acs.estlett.1c00375

Lee, W.L., McElroy, K.A., Armas, F., Imakaev, M., Gu, X., Duvallet, C., Chandra, F., Chen, H., Leifels, M., Mendola, S., Floyd-O’Sullivan, R., Powell, M.M., Wilson, S.T., Wu, F., Xiao, A., Moniz, K., Ghaeli, N., Matus, M., Thompson, J., Alm, E.J., 2021c. Quantitative detection of SARS-CoV-2 B.1.1.7 variant in wastewater by allele-specific RT-qPCR. medRxiv 2021.03.28.21254404. https://doi.org/10.1101/2021.03.28.21254404

Medema, G., Heijnen, L., Elsinga, G., Italiaander, R., Brouwer, A., 2020. Presence of SARS-Coronavirus-2 RNA in Sewage and Correlation with Reported COVID-19 Prevalence in the Early Stage of the Epidemic in The Netherlands. Environ. Sci. Technol. Lett. https://doi.org/10.1021/acs.estlett.0c00357

Napit, R., Manandhar, P., Chaudhary, A., Shrestha, B., Poudel, A., Raut, R., Pradhan, S., Raut, S., Mathema, S., Rajbhandari, R., Dixit, S., Schwind, J.S., Johnson, C.K., Mazet, J.K., Karmacharya, D., 2021. Rapid genomic surveillance of SARS-CoV-2 in a dense urban community using environmental (sewage) samples. medRxiv 2021.03.29.21254053. https://doi.org/10.1101/2021.03.29.21254053

Pal, M., Berhanu, G., Desalegn, C., Kandi, V., 2020. Severe Acute Respiratory Syndrome Coronavirus-2 (SARS-CoV-2): An Update. Cureus 12, e7423–e7423. https://doi.org/10.7759/cureus.7423

Petruska, J., Goodman, M.F., Boosalis, M.S., Sowers, L.C., Cheong, C., Tinoco, I.J., 1988. Comparison between DNA melting thermodynamics and DNA polymerase fidelity. Proc. Natl. Acad. Sci. U. S. A. 85, 6252–6256. https://doi.org/10.1073/pnas.85.17.6252

Polo, D., Quintela-Baluja, M., Corbishley, A., Jones, D.L., Singer, A.C., Graham, D.W., Romalde, J.L., 2020. Making waves: Wastewater-based epidemiology for COVID-19 – approaches and challenges for surveillance and prediction. Water Res. 186, 116404. https://doi.org/10.1016/j.watres.2020.116404

Pulliam, J.R.C., van Schalkwyk, C., Govender, N., von Gottberg, A., Cohen, C., Groome, M.J., Dushoff, J., Mlisana, K., Moultrie, H., 2021. Increased risk of SARS-CoV-2 reinfection associated with emergence of the Omicron variant in South Africa. medRxiv 2021.11.11.21266068. https://doi.org/10.1101/2021.11.11.21266068

Randazzo, W., Truchado, P., Cuevas-Ferrando, E., Simón, P., Allende, A., Sánchez, G., 2020. SARS-CoV-2 RNA in wastewater anticipated COVID-19 occurrence in a low prevalence area. Water Res. 181, 115942. https://doi.org/10.1016/j.watres.2020.115942

Thompson, J.R., Nancharaiah, Y. V, Gu, X., Lee, W.L., Rajal, V.B., Haines, M.B., Girones, R., Ng, L.C., Alm, E.J., Wuertz, S., 2020. Making waves: Wastewater surveillance of SARS-CoV-2 for population-based health management. Water Res. 184, 116181. https://doi.org/10.1016/j.watres.2020.116181

Van Poelvoorde, L.A.E., Delcourt, T., Coucke, W., Herman, P., De Keersmaecker, S.C.J., Saelens, X., Roosens, N., Vanneste, K., 2021. Strategy and performance evaluation of low-frequency variant calling for SARS-CoV-2 in wastewater using targeted deep Illumina sequencing. medRxiv 2021.07.02.21259923. https://doi.org/10.1101/2021.07.02.21259923

Wang, H., Miller, J.A., Verghese, M., Sibai, M., Solis, D., Mfuh, K.O., Jiang, B., Iwai, N., Mar, M., Huang, C., Yamamoto, F., Sahoo, M.K., Zehnder, J., Pinsky, B.A., 2021. Multiplex SARS-CoV-2 Genotyping PCR for Population-Level Variant Screening and Epidemiologic Surveillance. J. Clin. Microbiol. 59. https://doi.org/10.1101/2021.04.20.21255480

WHO, 2020. WHO Director-General’s opening remarks at the media briefing on COVID-19 −11 March 2020 [WWW Document]. URLhttps://www.who.int/director-general/speeches/detail/who-director-general-s-opening-remarks-at-the-media-briefing-on-covid-19---11-march-2020 (accessed 12.19.21).

WHO, 2021a. Tracking SARS-CoV-2 variants [WWW Document]. URL https://www.who.int/en/activities/tracking-SARS-CoV-2-variants/ (accessed 12.19.21).

WHO, 2021b. Enhancing Readiness for Omicron (B.1.1.529): Technical Brief and Priority Actions for Member States [WWW Document]. URL https://www.who.int/publications/m/item/enhancing-readiness-for-omicron-(b.1.1.529)-technical-brief-and-priority-actions-for-member-states (accessed 12.19.21).

Wu, D.Y., Ugozzoli, L., Pal, B.K., Wallace, R.B., 1989. Allele-specific enzymatic amplification of beta-globin genomic DNA for diagnosis of sickle cell anemia. Proc. Natl. Acad. Sci. U. S. A. 86, 2757–2760. https://doi.org/10.1073/pnas.86.8.2757

Yaniv, K., Ozer, E., Shagan, M., Lakkakula, S., Plotkin, N., Bhandarkar, N.S., Kushmaro, A., 2021. Direct RT-qPCR assay for SARS-CoV-2 variants of concern (Alpha, B.1.1.7 and Beta, B.1.351) detection and quantification in wastewater. Environ. Res. 201, 111653. https://doi.org/10.1016/j.envres.2021.111653

Wu, F., Xiao, A., Zhang, J., Moniz, K., Endo, N., Armas, F., Bonneau, R., Brown, M.A., Bushman, M., Chai, P.R., Duvallet, C., Erickson, T.B., Foppe, K., Ghaeli, N., Gu, X., Hanage, W.P., Huang, K.H., Lee, W.L., Matus, M., McElroy, K.A., Nagler, J., Rhode, S.F., Santillana, M., Tucker, J.A., Wuertz, S., Zhao, S., Thompson, J., Alm, E.J., 2022. SARS-CoV-2 RNA concentrations in wastewater foreshadow dynamics and clinical presentation of new COVID-19 cases. Sci. Total Environ. 805, 150121. https://doi.org/10.1016/j.scitotenv.2021.150121

Wu, F., Xiao, A., Zhang, J., Moniz, K., Endo, N., Armas, F., Bushman, M., Chai, P.R., Duvallet, C., Erickson, T.B., Foppe, K., Ghaeli, N., Gu, X., Hanage, W.P., Huang, K.H., Lee, W.L., Matus, M., McElroy, K.A., Rhode, S.F., Wuertz, S., Thompson, J., Alm, E.J., 2021. Wastewater Surveillance of SARS-CoV-2 across 40 U.S. states. medRxiv 2021.03.10.21253235. https://doi.org/10.1101/2021.03.10.21253235

Wu, F., Zhang, J., Xiao, A., Gu, X., Lee, W.L., Armas, F., Kauffman, K., Hanage, W., Matus, M., Ghaeli, N., Endo, N., Duvallet, C., Poyet, M., Moniz, K., Washburne, A.D., Erickson, T.B., Chai, P.R., Thompson, J., Alm, E.J., 2020. SARS-CoV-2 Titers in Wastewater Are Higher than Expected from Clinically Confirmed Cases. mSystems 5, e00614–20. https://doi.org/10.1128/mSystems.00614-20

Zhou, P., Yang, X.-L., Wang, X.-G., Hu, B., Zhang, L., Zhang, W., Si, H.-R., Zhu, Y., Li, B., Huang, C.-L., Chen, H.-D., Chen, J., Luo, Y., Guo, H., Jiang, R.-D., Liu, M.-Q., Chen, Y., Shen, X.-R., Wang, X., Zheng, X.-S., Zhao, K., Chen, Q.-J., Deng, F., Liu, L.-L., Yan, B., Zhan, F.-X., Wang, Y.-Y., Xiao, G.-F., Shi, Z.-L., 2020. A pneumonia outbreak associated with a new coronavirus of probable bat origin. Nature 579, 270–273. https://doi.org/10.1038/s41586-020-2012-7

